# Distinct microRNA expression signatures of primary and secondary central nervous system lymphomas

**DOI:** 10.1101/2021.02.05.21249862

**Authors:** Endre Sebestyén, Ákos Nagy, Dóra Marosvári, Hajnalka Rajnai, Béla Kajtár, Beáta Deák, András Matolcsy, Sebastian Brandner, James Storhoff, Ning Chen, Attila G. Bagó, Csaba Bödör, Lilla Reiniger

## Abstract

Central nervous system (CNS) lymphoma is a rare and aggressive non-Hodgkin lymphoma that might arise in the CNS (primary CNS lymphoma, PCNSL) or disseminates from a systemic lymphoma to the CNS (secondary CNS lymphoma, SCNSL). Dysregulated expression of microRNAs (miRNAs) is associated with various pathological processes and miRNA expression patterns may have diagnostic, prognostic and therapeutic implications. However, miRNA expression is understudied in CNS lymphomas. Here, we performed expression analysis of 798 miRNAs in 73 CNS lymphoma samples using the NanoString platform, followed by a detailed statistical analysis to identify potential novel biomarkers characterizing subgroups and to examine differences based on their primary and secondary nature, molecular subtype, mutational patterns and survival. We describe the general expression patterns of miRNAs across CNS lymphoma samples and identified 31 differentially expressed miRNAs between primary and secondary groups. Additionally, we identified 7 more miRNAs associated with a molecular subtype and 25 associated with mutation status. Using unsupervised clustering methods, we defined a small but distinct primary CNS lymphoma subgroup, with characteristically different expression patterns compared to the rest of the cases. Finally, we identified differentially regulated pathways in the above comparisons and assessed the utility of miRNA expression patterns in predicting survival. Our study identifies a novel CNS lymphoma subgroup defined by distinct miRNAs, proves the importance of specific miRNAs and pathways in their pathogenesis, and provides the basis for future research.

## Background

Central nervous system (CNS) lymphoma is a rare and aggressive non-Hodgkin lymphoma that either arises in the CNS structures (primary CNS lymphoma, PCNSL) or disseminates from a systemic lymphoma to the CNS (secondary CNS lymphoma, SCNSL). Histologically, it predominantly manifests as a diffuse large B-cell lymphoma (DLBCL). Effective treatment of CNS lymphomas remains a significant challenge as their molecular pathogenesis is not well understood [1-4].

Both from a therapeutic and prognostic point of view, it is becoming increasingly important to precisely define the molecular subtype of DLBCLs into germinal center B-cell (GC) type, activated B-cell (ABC) type or “unclassified” (UC) cases, as described by Alizadeh *et al*. [5]. Patients in the ABC-type DLBCL group show an inferior outcome [5, 6] compared to the other types. The fundamental difference in biology including oncogenic pathways and mutation targets between GC- and ABC-type DLBCLs is also reflected in the different efficacy of novel targeted therapies between these subgroups [7, 8]. A more precise, gene expression-based molecular subtype assignment can be achieved from formalin-fixed paraffin embedded (FFPE) tissue using the NanoString Lymphoma Subtyping Test (LST) assay (NanoString Technologies, Inc., Seattle, USA) compared to the standard immunohistochemical (IHC) methods. The NanoString assay also demonstrates a better concordance with the gold-standard Affymetrix approach [9].

The discovery of microRNAs (miRNAs, miRs) has opened a new field for unraveling and therapeutically targeting diseases. These small non-coding RNAs regulate diverse biological processes through post-transcriptional gene expression modulation. Based on their seed sequence, miRNAs bind to multiple target mRNAs, thereby promoting their degradation or inhibiting translation [10-12]. Dysregulated expression of miRNAs is associated with a myriad of pathological processes including hematological malignancies [13, 14], and distinct miRNA expression patterns may also have diagnostic, prognostic and therapeutic implications [15-19]. As miRNAs remain relatively well preserved in archival FFPE tissue specimens, they are readily available as a valuable source of information in cancer tissues [20-22]. For the quantification of miRNA transcripts in FFPE samples, the NanoString nCounter technology is a preferable choice over quantitative reverse transcription polymerase chain reaction (RT-PCR) [23, 24], with a high reproducibility similar to other platforms [25, 26]. The NanoString assay also performs well in relative quantification studies [27].

MiRNA expression of PCNSL has been studied using different methods such as RT-PCR [28-38], microarray [36, 39], *in situ* hybridization [37, 38], next-generation sequencing (NGS) [35] and NanoString [38] technologies on various tissue types including brain biopsy specimens (FFPE [32, 37, 39] or fresh [36]), cerebrospinal fluid (CSF) [28-31, 33, 38] and peripheral blood [35], serum [34] or plasma [33]. It has been shown and further confirmed that the combined detection of miR-21, miR-19b and miR-92a in CSF allowed a reliable diagnosis of PCNSL. Moreover, these miRNAs emerge as promising tools in treatment monitoring and follow-up [28, 29], similarly to U2 small nuclear RNA fragments [30]. In addition, plasma miR-21 may serve as a diagnostic [33], and serum miR-21 both as a diagnostic and prognostic marker for PCNSL [34]. Other miRNAs with prognostic value in PCNSL include miR-151a-5p and miR-151b, together with 10 additional miRs [35]. CSF levels of miRlzl21 may have potential as a predictor of chemotherapeutic effect [33]. Measuring miR-30c in the CSF can differentiate between PCNSL and SCNSL, as elevated levels of miR-30c have pathobiological significance in SCNSL [31]. A recent microarray study found a couple of miRNAs with positive or negative prognostic significance in PCNSL [36]. It has also been demonstrated, that PCNSL shows different miRNA expression profiles compared with nodal or testicular DLBCL [32, 37, 39].

In this study, we performed expression profiling of 798 human miRNAs in 73 FFPE brain biopsy samples of primary and secondary CNS lymphomas using the NanoString platform, followed by a bioinformatics analysis to reveal changing expression signatures. We aimed to identify potential novel biomarkers characterizing subgroups among brain lymphomas, as well as to examine differences based on their primary and secondary nature, molecular subtype, mutational patterns and survival.

## Methods

### Sample collection and patient information

FFPE brain biopsy specimens of 64 patients with PCNSL and 9 patients with SCNSL were analyzed in this study. Tissue samples were obtained from three centers: (i) 1^st^ Department of Pathology and Experimental Cancer Research, Semmelweis University, Budapest, Hungary; (ii) Department of Pathology, University of Pécs, Pécs, Hungary and (iii) Division of Neuropathology, The National Hospital for Neurology and Neurosurgery, University College London Hospitals, United Kingdom, through the UK Brain Archive Information Network (BRAIN UK). Permissions to use the archived tissue have been obtained from the Local Ethical Committee (TUKEB-1552012) and from BRAIN UK (Ref.: 16/018), and the study was conducted in accordance with the Declaration of Helsinki.

Information on the molecular subtypes determined by the Research Use Only version of the NanoString LST-assay (NanoString Technologies, Inc., Seattle, USA) and on the mutational status identified by ultra-deep NGS of 14 target genes as described previously [40] are summarized in Supplementary file 1, together with clinical and survival data. Survival data was available in 54 PCNSL and 9 SCNSL cases. The PCNSL cases consisted of 78.1% (50/64) ABC, 15.6% (10/64) GC and 6.3% (4/64) UC molecular subtypes. Among the SCNSL cases, 44.4% (4/9) were classified as ABC- and 55.6% (5/9) as GC-subtypes (Figure 1A).

**Figure 1.**
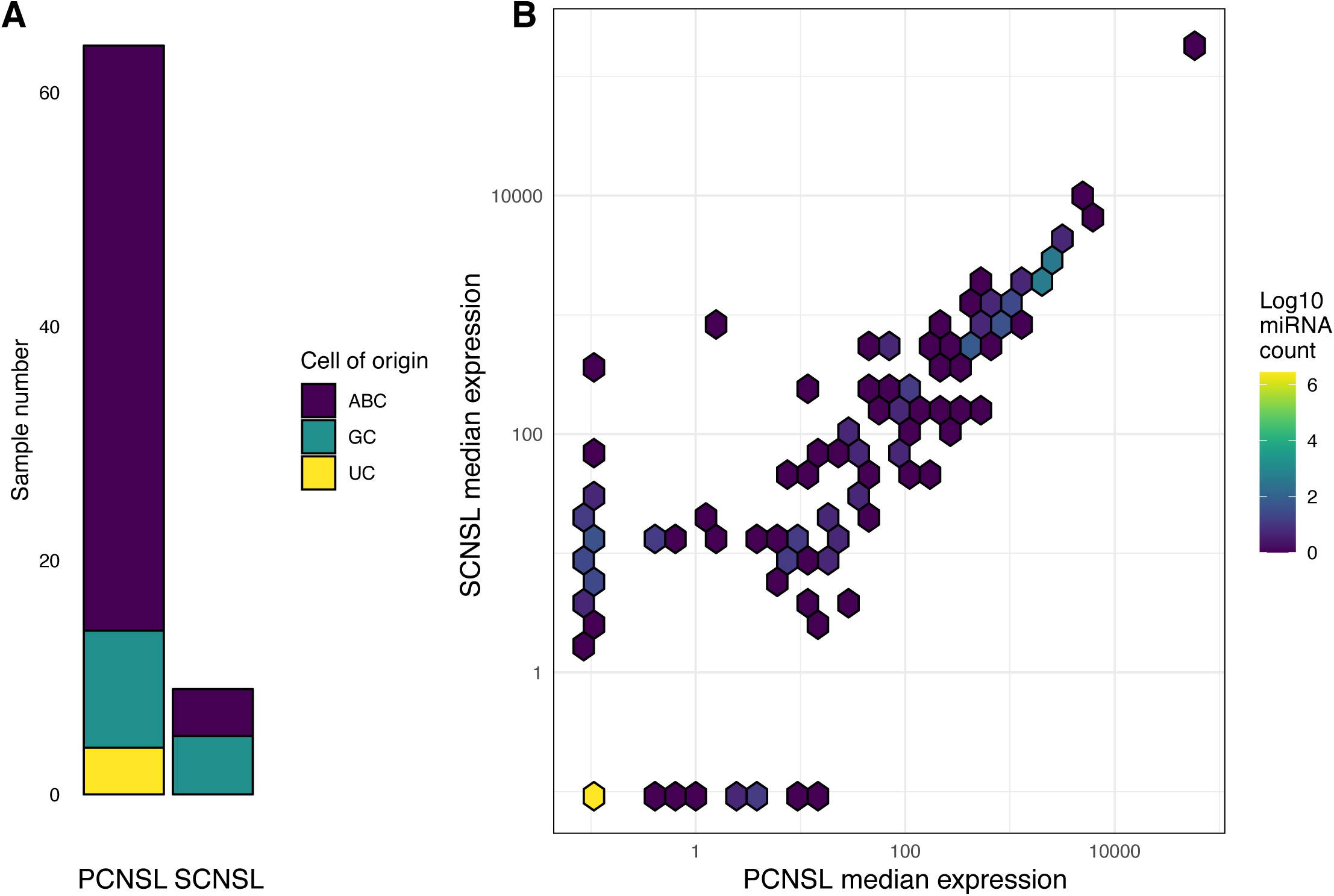
Sample characteristics and general expression patterns. **a**) Total number of primary and secondary central nervous system lymphoma samples categorized by molecular subtypes. **b**) Median normalized expression of miRNAs in PCNSL (x-axis) and SCNSL (y-axis) samples. Both axes are log_10_ based, and the hexagon color scale shows the number of miRNAs falling into a particular median expression range. As can be seen on the plot (bright yellow hexagon in the bottom left corner), a large number of miRNAs had a 0 or near 0 expression in both sample types. The Spearman correlation of PCNSL and SCNSL values is 0.89.

### MicroRNA profiling using the NanoString platform

RNA isolation from 64 PCNSL and 9 SCNSL samples was performed using the RecoverAll™ kit (Life Technologies/Ambion, Inc, Foster City USA) according to the manufacturer’s instructions. Approximately 100 ng of total RNA from each sample was analyzed using the Human v3 miRNA expression assay kit, following manufacturer’s instructions (NanoString Technologies, Inc., Seattle, USA). The Human v3 miRNA assay covers 98% of miRNA sequences found in miRBase v22, including 798 probes that recognize human miRNAs and 29 assay control probes. Raw data were pulled from digital analyzer and imported into nSolver4.0 (NanoString™) for data quality check. Data sets that passed all QC checks (including imaging quality, binding density, positive control linearity and limit of detection) were exported as a .csv file for downstream statistical analysis. Raw miRNA counts for all samples are available in the GSE162956 dataset at NCBI GEO.

### NanoString data normalization

Raw NanoString read counts and clinical information was imported into the R statistical environment (version 3.5.1). We discarded human miRNAs with extremely low expression (read count <= 1 in more than 60 samples) before normalization, resulting in 781 human miRNAs, besides the control probes and housekeeping genes. Data normalization was done with the NanoStringNorm package (version 1.2.1) [41] using the following options: CodeCount = “geo.mean”, Background = “mean.2sd”, SampleContent = “housekeeping.geo.mean”, OtherNorm = “quantile”. We used the ath, cel, osa and NEG miRNA classes as negative controls, the POS class as positive controls, the *ACTB, B2M, GAPDH, RPL19* and *RPLP0* genes as housekeeping genes, and everything else as endogenous miRNAs. During the initial data assessment, we considered those miRNAs expressed, that had a larger normalized expression value than the lowest positive control (Addt 2). Additionally, we calculated the minimum, maximum and average expression of all miRNAs across all samples, together with the number of samples where a miRNA was expressed across all samples using the normalized expression values. Finally, we calculated the minimum, maximum, average and median values for the PCNSL and SCNSL samples separately. Based on the median expression values we ranked the miRNAs and categorized them into 15 separate expression groups.

### Differential expression analysis

We used the limma package (version 3.38.3) for differential expression [42]. We included the primary or secondary category, the molecular subtype, origin of sample (institute or department), degradation time and NanoString scan dates in the linear model as covariates during differential expression analysis. We converted the normalized read counts with the voom [43] function of limma, fitted the linear model with the lmFit function and calculated p-values with the eBayes function [44]. The raw p-values were false discovery rate (FDR) adjusted using the Benjamini-Hochberg method. We considered miRNAs differentially expressed with FDR adjusted p-value < 0.05 and absolute log2 fold change between conditions > 1 (Supplementary file 3).

We carried out a number of comparisons between different sample groups. First, we calculated differentially expressed miRNAs between the primary and secondary samples. Additionally, we calculated differentially expressed miRNAs between the different molecular subtypes, within the primary samples. This included the GC vs ABC, UC vs ABC and GC vs UC comparisons. Finally, we calculated differences between the different sample groups stratified by mutation status, using the *CARD11, CCND3, CD79B, CSMD2, CSMD3, IRF4, KMT2D, MYC, MYD88, PAX5, PIM1, PRDM1* and *TP53* genes (Supplementary file 3).

### Principal component analysis and unsupervised clustering

During the principal component analysis, we used the normalized and voom transformed data, additionally removing the potential batch effects of the institute, scan date and degradation time with the removeBatchEffect function of the limma package. We calculated the principal components with the prcomp R function with the scale. parameter set to TRUE, and plotted the first and second principal components.

During the binary hierarchical clustering of the samples, we used the voom transformed, normalized read counts of all endogenous miRNAs, after removing the potential batch effects of the institute, scan date and degradation time with the removeBatchEffect function of the limma package. First, we binarized the expression values [45] with a cut-off of 12. All miRNAs below this threshold were considered not expressed, while the rest was considered expressed. Using the expressed/not expressed status of the miRNAs across all samples, we calculated sample distances using the dist function from R, with the binary method, and did a hierarchical clustering using the hclust function, with the ward.D2 method. We manually defined a “small” and a “big” cluster of samples (Supplementary file 4), and carried out a Fisher-test, using a 2×2 contingency table (expressed/not expressed and small/big cluster) to check for miRNAs whose expression is associated with the small and big clusters (Supplementary file 5). The Fisher-test p-values were FDR adjusted using the Benjamini-Hochberg method and miRNAs with FDR adjusted p-value < 0.05 were considered associated with the cluster type. We repeated the binary hierarchical clustering using only the primary samples without changing any other parameter.

Additionally, we performed a k-means clustering on the data, where we also used the voom transformed, normalized read counts of all endogenous miRNAs after removing the potential batch effects of the institute, scan date and degradation time with the removeBatchEffect function of the limma package. We used kmeans function of R, with the iter.max = 1000 parameter and setting the cluster number to 4. We clustered both the samples and the miRNAs. During data visualization we dropped the largest miRNA cluster as it mainly contained miRNAs that were not expressed in most samples or had a very low expression level. We repeated the k-means clustering using only the primary samples without changing any other parameter.

### miRNA pathway enrichment analysis

We used the miRNet database (accessed and downloaded on 2019-08-29) to collect putative targets of the differentially expressed miRNAs [46]. We downloaded all gene and lncRNA targets of these miRNAs from bone-marrow or brain tissue-based experiments. We considered a gene up- or downregulated if at least two of its putative regulators from miRNet were differentially expressed in the opposite direction in a specific tissue type. For example, the AGO1 gene was considered downregulated in the secondary vs primary comparison, as 7 of its regulatory miRNAs defined in miRNet were upregulated in the secondary vs primary comparison based on the NanoString analysis. Based on this filtering criteria, only bone marrow-based mRNA interactions were considered for further downstream analysis.

Additionally, we downloaded version 7 of the MSigDB gene sets (H, C2 – C7) [47] and did a Fisher-test using a 2×2 contingency table as follows: genes were either part of a gene collection or not, and based on the miRNet analysis they were putatively differentially expressed or not. The Fisher-test p-values were FDR adjusted using the Benjamini-Hochberg method and gene collections with FDR adjusted p-value < 0.05 were considered enriched in a specific comparison (secondary vs primary, etc) (Supplementary file 6).

### Survival analysis

Survival data was available for 63 patients with the overall survival in months, last follow-up date, and survival status. We used only the 54 primary CNS lymphoma cases during this analysis, and stratified patients according to the binary expression status of the investigated miRNAs, or the big/small cluster classification. We used the survival package (version 2.44-1.1) [48] to create Surv objects in R and calculated log-rank test based p-values using the survminer (version 0.4.6) [49] package. The log-rank p-values were FDR adjusted using the Benjamini-Hochberg method (Supplementary file 7).

### Validation of NanoString miRNA expression data

10 miRNAs with significant differential expression among subgroups were selected (Supplementary file 8 and 9) based on NanoString expression data. Additionally, 3 miRNAs with stable expression in the cohort were used as endogenous control.

#### Validation with droplet digital PCR

The selected samples were individually reverse-transcribed with the TaqMan™ MicroRNA Reverse Transcription Kit (Applied Biosystems, USA) following manufacturer’s instructions. Hundred ng miRNA from each selected sample was reverse-transcribed with specific TaqMan™ miRNA assays (Applied Biosystems, USA) in ProFlex Thermal Cyclers (Applied Biosystems, USA) using the following parameter values: 16°C for 30 min, 42°C for 30 and 85°C for 5 min. cDNAs were further diluted with 50 ul nuclease-free water to obtain a final volume of 77 ul. 10 ul diluted cDNA sample were used in the subsequent droplet digital PCR (ddPCR) reactions with 1 ul of the specific TaqMan™ miRNA assays and 11 ul of ddPCR Supermix for Probes (No dUTP) (Bio-Rad Laboratories, USA). Following droplet generation, target miRNAs were amplified in a C1000 Touch Thermal Cycler (Bio-Rad Laboratories, USA) with parameters as follows: 10 min at 95°C for enzyme activation, followed by 40 cycles of denaturation at 94°C for 30 s and 1 min annealing/extension at 55°C, enzyme deactivation step set at 98°C for 10 min, and a final hold step at 4°C for an infinite time period. Results were analyzed using the QuantaSoft software (version 1.7; Bio-Rad, USA). All ddPCR reactions were performed with the detection of adequate events (>10000 droplets per sample). Results were determined in copy number per ul.

#### Validation with quantitative RT-PCR

Further NanoString miRNA expression data validation was completed with quantitative RT-PCR. Reverse Transcription was performed as described above with the exception that each miRNA target was individually reverse-transcribed with an endogenous control miRNA. Five ul of the cDNA samples were amplified in duplicates using 1.5 ul of the target and endogenous control specific TaqMan™ miRNA assays with 7.7 ul 2x TaqMan puffer (Applied Biosystems, USA) and 10.5 ul nuclease-free water on a Quantstudio 3 Real-Time PCR system (Applied Biosystems, USA). Ct values for sample target and endogenous miRNAs were determined as the average Ct value of the duplicates. Δ-Ct values were obtained extracting Ct values of endogenous control miRNAs from the Ct values of target miRNAs, which were subsequently used for statistical analysis.

## Results

### General miRNA expression patterns in CNS lymphomas

The NanoString panel contained 798 endogenous, 5 housekeeping genes (*ACTB, B2M, GAPDH, RPL19, RPLP0*), 9 positive and 16 negative controls. Based on the normalized expression values, almost half of the investigated miRNAs (381) were not expressed in any sample, and 54 were expressed only in one. On the other hand, 12 miRNAs were expressed in 72 and 2 miRNAs in all 73 samples (Supplementary file 2).

We compared the median expression of endogenous miRNAs between PCNSL and SCNSL samples (Figure 1B). miRNAs with a non-zero median expression in both groups were highly correlated (Spearman correlation 0.89). We found 676 and 661 miRNAs in PCNSL and SCNSL respectively, that had rank in the bottom 10 % based on their median expression. On the other hand, hsa-miR-4454+hsa-miR-7975 is an extreme outlier, with a larger median expression than most of the positive controls. We noticed that the hsa-miR-4454 expression detected by NanoString might be confounded by the expression of tRNA^His^, as the miRNA is identical to the 3’ end of this tRNA [50].

### Differential miRNA expression between primary and secondary CNS lymphomas and molecular subtypes

We analyzed differential expression patterns of the miRNAs between PCNSL and SCNSL cases and different molecular subtypes using the limma R package, after removing known confounding factors. With an absolute log_2_ fold change > 1 and FDR value < 0.05 cut-off, we found 31, 4 and 3 differentially expressed miRNAs between PCNSL and SCNSL cases, GC and ABC subtypes, and UC and ABC subtypes, respectively (Figure 2A, Table 1 and Supplementary file 3). Twenty-eight miRNAs showed increased and three showed decreased expression in SCNSL compared to PCNSL. Three miRNAs showed lower and one miRNA higher expression in GC compared to ABC cases. All three differentially expressed miRNAs showed lower levels in UC compared to ABC subtypes (Supplementary file 3).

**Table 1.**
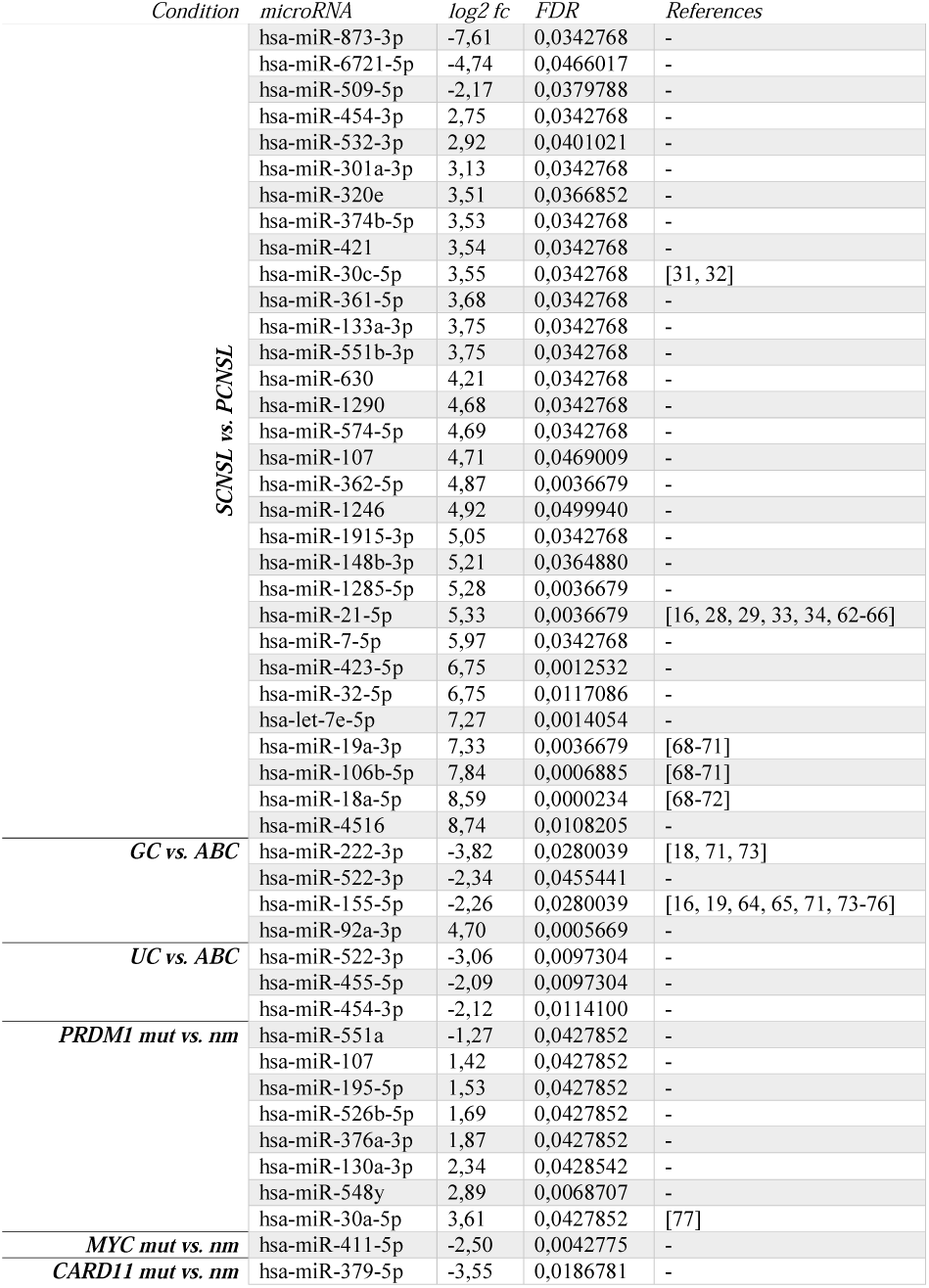
Differentially expressed miRNAs between primary and secondary central nervous system lymphomas, molecular subtypes and in association to mutation status. The columns show the tested condition, the log_2_ fold-change, the FDR and the literature references if known. Abbreviations: ABC, activated B-cell type; GC, germinal center B-cell type; mut, mutated; nm, non-mutated; PCNSL, primary central nervous system lymphoma; SCNSL, secondary central nervous system lymphoma; UC, unclassified; vs., versus.

**Figure 2.**
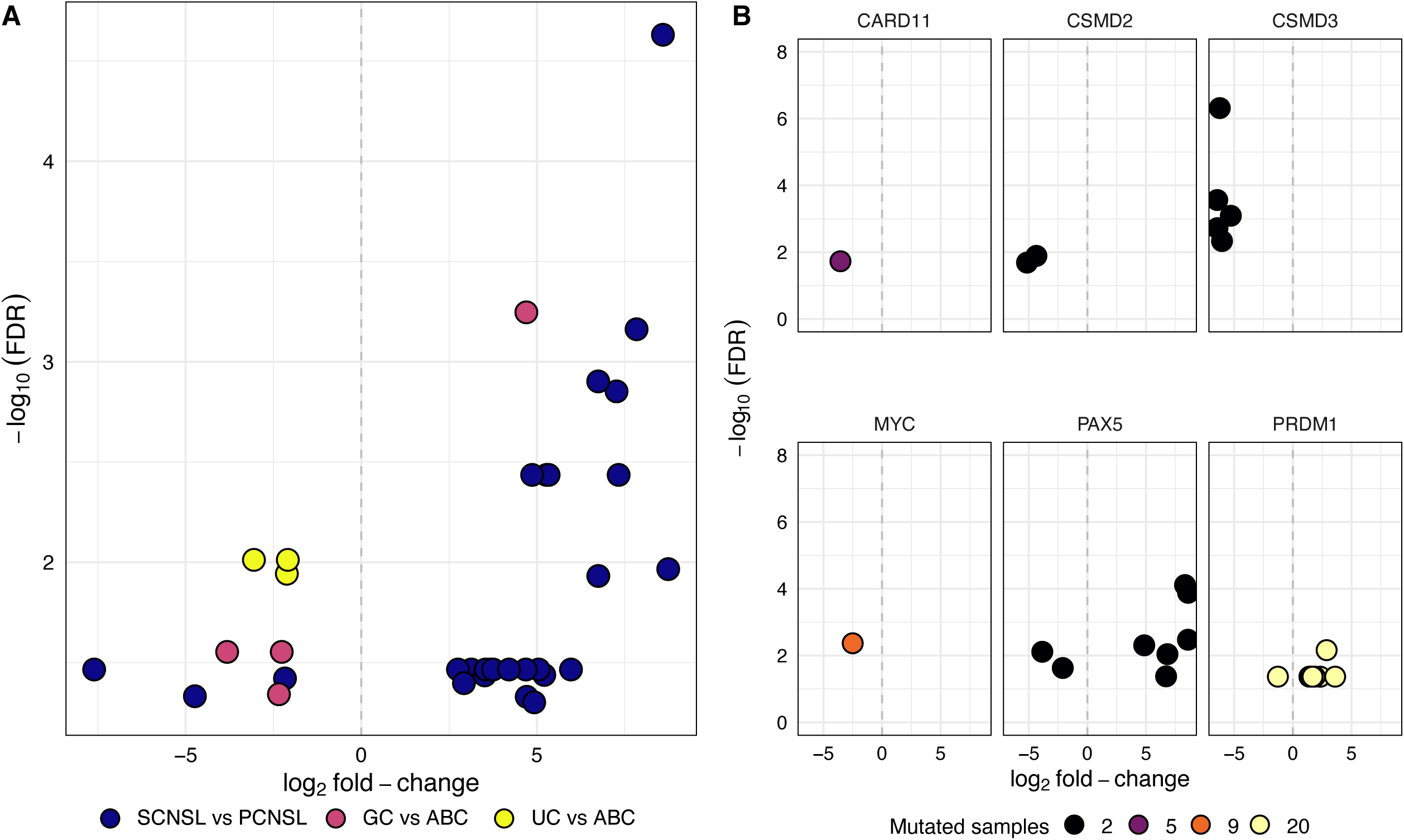
Differential expression analysis between sample groups. **a**) Volcano-plot of differential expression results comparing secondary and primary samples, the GC and ABC or UC and ABC sample groups. The x-axis shows the log_2_ fold change of a specific miRNA, while the y-axis shows the -log_10_ transformed FDR corrected p-value. **b**) Volcano-plot of differential expression results comparing mutated and non-mutated samples for a specific gene, where the color of the dots correlates with the number of mutated samples. As in panel **a**) x-axis shows the log_2_ fold change of a specific miRNA, while the y-axis shows the -log_10_ transformed FDR corrected p-value.

### miRNA expression profile association to mutation status

We assessed whether miRNA expression profiles were associated with the mutation status of specific genes. We found 8 differentially expressed miRNAs associated with the mutation status of the *PRDM1* gene, and one miRNA associated with the mutation status of *MYC* and *CARD11* genes (Figure 2B, Table 1 and Supplementary file 3).

We also identified 8, 5 and 2 differentially expressed miRNAs in cases with *PAX5, CSMD3* and *CSMD2* mutations. However, there were only two samples harboring mutation in each of these genes, therefore, the results must be considered with caution (Supplementary file 3).

### Unsupervised clustering of miRNA expression data

We checked if the data grouped according to disease characteristics using principal component analysis (PCA). The PCA results did not indicate the presence of distinct groups based on disease subtype (primary or secondary) or molecular subtype (Figure 3). Additionally, the samples do not cluster based on RNA isolation group, place of origin, scanning date, degradation time, % tumor content, age or sex (Supplementary Figure 1 – 8). As we could not define any specific grouping of samples based on this analysis, we investigated the miRNA expression patterns further, using additional clustering methods.

**Figure 3.**
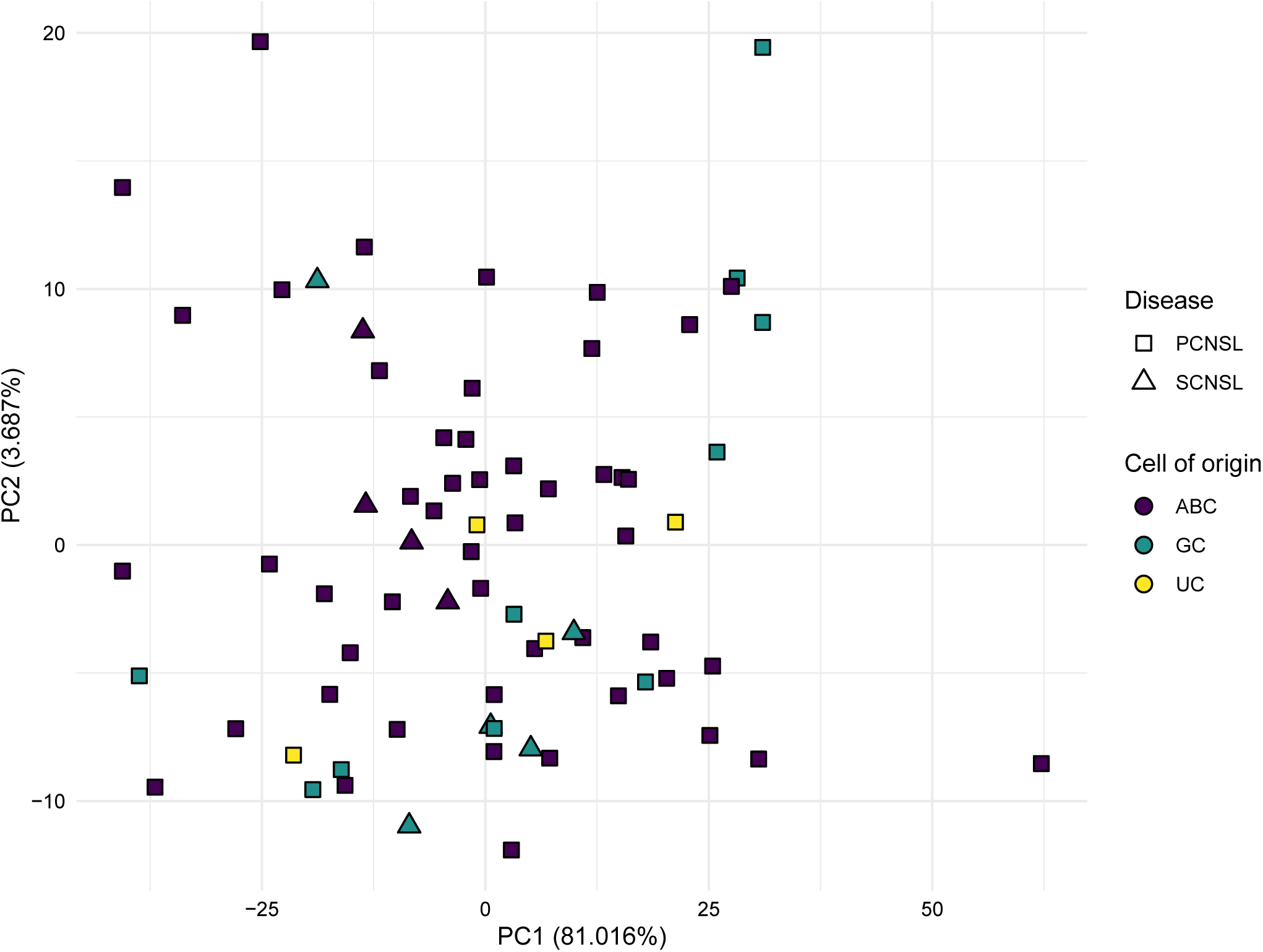
Principal component analysis of miRNA expression patterns of all samples, after removing potential batch effects from the normalized and voom transformed data. The shape of the points shows the primary or secondary disease category, while the color corresponds to the molecular subtype.

First, we did a binary clustering of the data (Figure 4), where we only considered the expressed/not expressed status of miRNAs. Based on the binary clustering, two sample groups become apparent. A small group, consisting of 8 samples are clearly distinct from the rest of the cohort (Supplementary file 4). The binary expression patterns of this group are markedly different from the larger group. For example, the hsa-miR-93-5p and hsa-let-7d-5p miRNAs were not expressed in any samples from the small cluster, while the hsa-miR-181a-5p miRNA was expressed only in one sample. These miRNAs were expressed only in a small fraction of the large cluster. Considering these expression patterns, we carried out a systematic analysis and validated if the expression of specific miRNAs is significantly associated with the small and large groups using a Fisher test. Based on the test results, we found 19 miRNAs, whose expression patterns are associated with the clusters (Supplementary file 5). Interestingly, we only found miRNAs, whose lack of expression was associated with the small cluster, and did not find any, whose presence of expression was associated with the same cluster. Based on these patterns, we initially thought that the small cluster might be the result of a particularly bad quality sample group or strong technical bias. However, based on the normalized, batch-corrected PCA analysis (Supplementary Figures 1-8) of the expression data, this does not seem to be true. The cluster remains even after correcting for all known technical biases, and also does not seem to be associated with age or sex.

**Figure 4.**
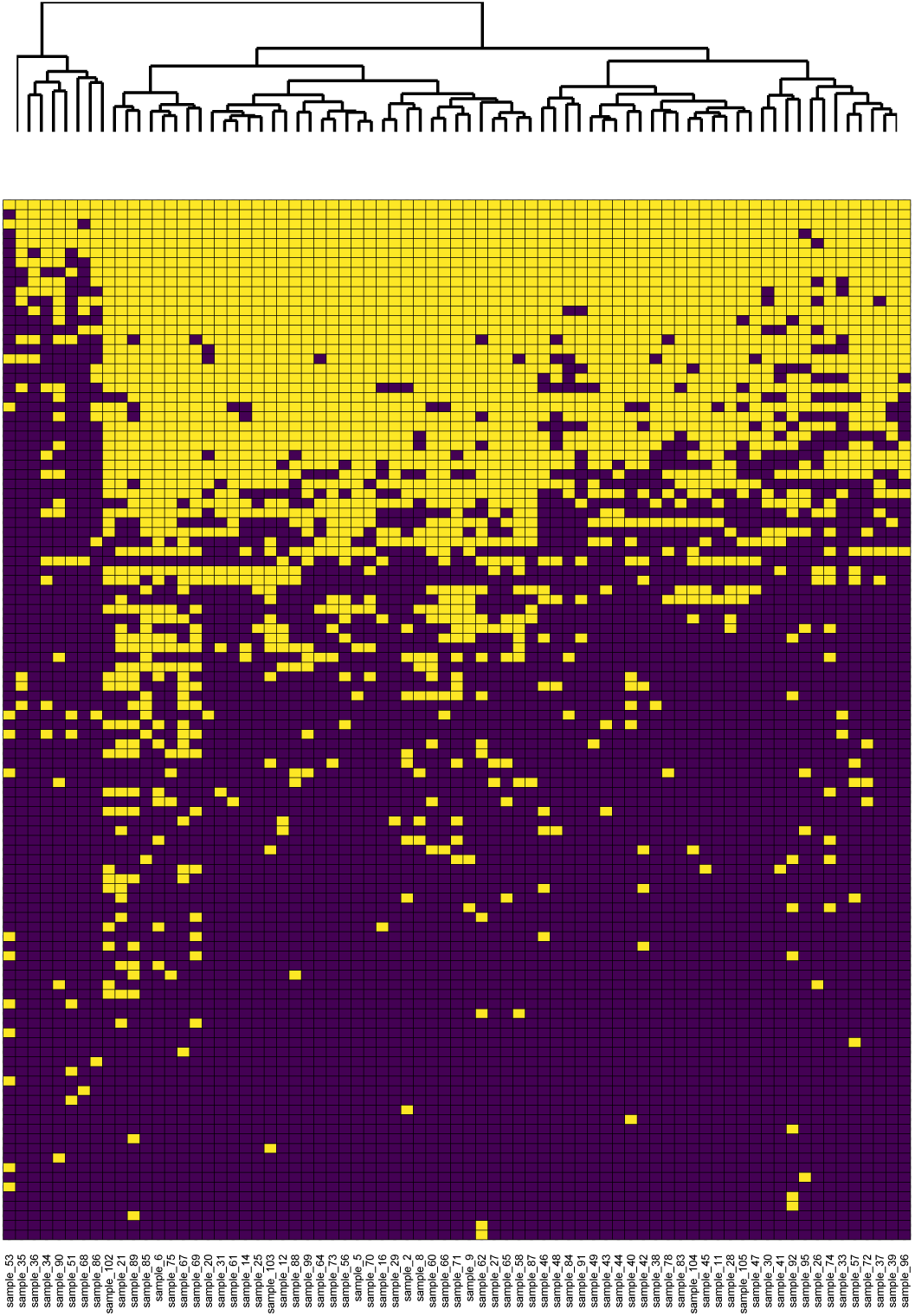
Unsupervised clustering of samples using binary (ON/OFF) miRNA expression patterns. The heatmap shows the binary expression pattern of miRNAs that were considered expressed in at least one sample. Columns correspond to samples, while rows correspond to a specific miRNA. Yellow tiles show expressed, purple tiles show non-expressed miRNAs. On top of the heatmap a hierarchical clustering tree shows the relationship between samples. miRNAs that were not expressed in any of the samples are not shown.

After the binary clustering analysis, we repeated sample clustering using a k-means based method and instead of the binary expression data, we used normalized expression values (Figure 5). Based on visual inspection, we decided to use 4 clusters for the k-means algorithm, both for the sample and the miRNA level analysis. Interestingly, the second largest k-means sample cluster (SCluster1) consisting of 10 samples, partially overlaps with the “small” cluster defined in the binary expression analysis (Supplementary file 4). In summary, both the binary expression clustering and the k-means clustering defined a small set of samples with markedly distinct miRNA expression patterns from the rest.

**Figure 5.**
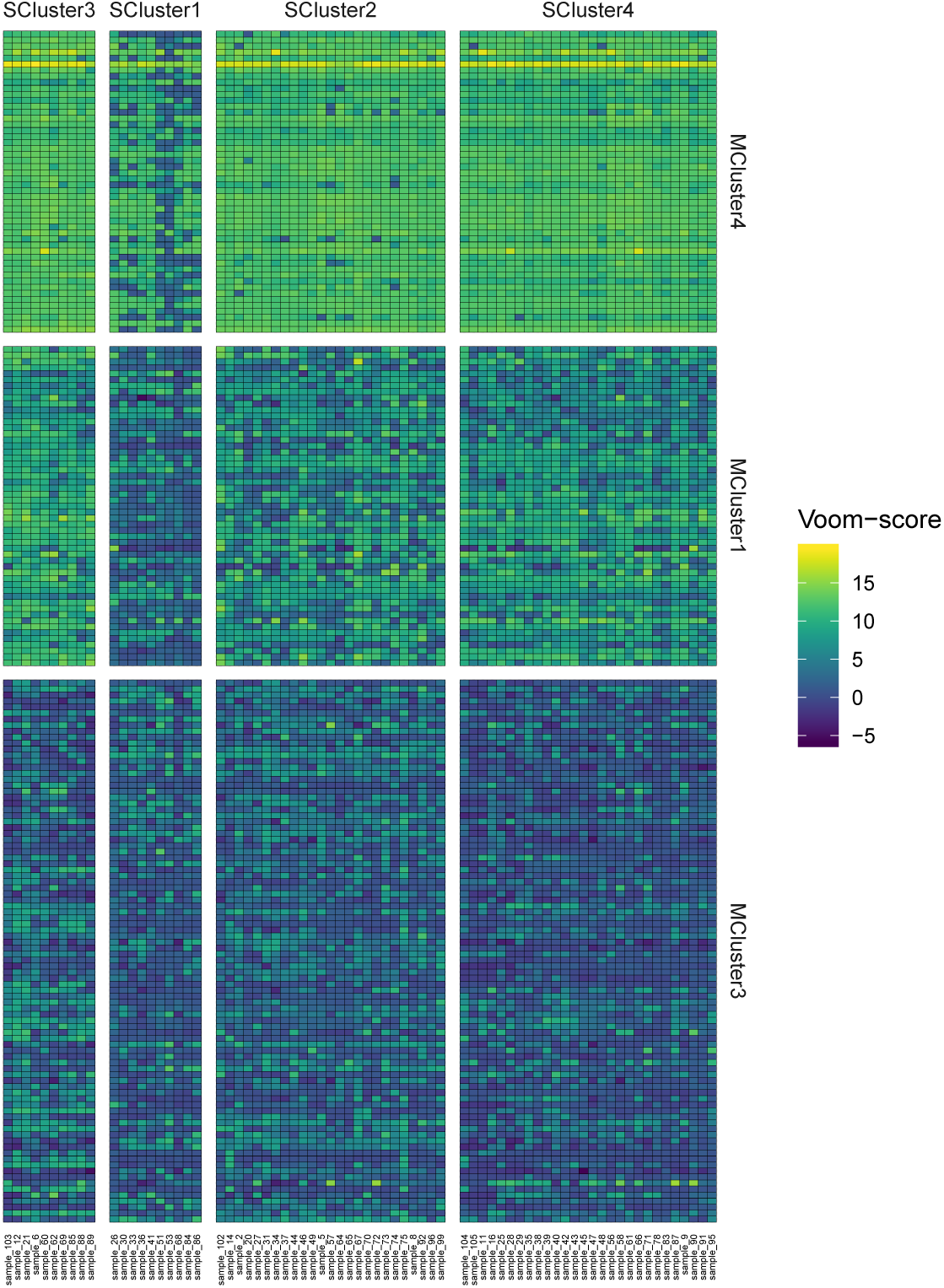
Unsupervised clustering of samples using normalized miRNA expression patterns. The heatmap shows the k-means clustering of miRNAs after normalization, voom transformation and batch effect removal. Columns correspond to samples, while rows correspond to a specific miRNA. Heatmap colors show expression intensity. We removed the largest miRNA cluster (Mcluster2) from the visualization as it contained mainly miRNAs with zero or very low expression.

After inspecting the miRNA k-means clusters, we noticed that the expression of miRNAs in the second largest cluster (MCluster1) in SCluster1 is low for most of the samples. We checked the overlap of MCluster1 miRNAs with the 19 miRNAs that had significantly different binary expression in the previous analysis. All of the 19 miRNAs were present in the new k-means based cluster. Therefore, MCluster1 contains those miRNAs whose expression pattern defines the small sample cluster seen in all of the above analysis.

Repeating both analyses using only PCNSL samples lead to similar results, with the only difference being that the “small” binary cluster contained an additional sample (sample no. 33) (Supplementary Figures 9 and 10).

### Pathway enrichment of differentially expressed miRNAs

After the differential expression analysis, we asked if specific pathways might be up- or downregulated in a specific comparison. Assuming that the differential regulation of a specific miRNA will lead to the differential regulation of its target mRNAs in the opposite direction, we carried out a pathway enrichment analysis, based on validated miRNA-mRNA interactions from the miRNET database. Due to the low number of differentially expressed miRNAs, we were able to do this analysis only in the case of SCNSL vs PCNSL and *PRDM1* mutated vs non-mutated comparisons. Focusing on the hallmarks (H) gene sets of MSigDB, we found several pathways that are putatively downregulated in these cases (Figure 6 and Supplementary file 6). The G2M checkpoint, PI3K-AKT-MTOR signaling, TGF-beta signaling pathways, MYC target and androgen response genes were downregulated in both comparisons besides a number of other pathways or gene sets. Additionally, in the SCNSL vs PCNSL comparison TNF-alpha signaling, apoptosis and UV response genes, the P53 pathway and E2F target genes were also downregulated.

**Figure 6.**
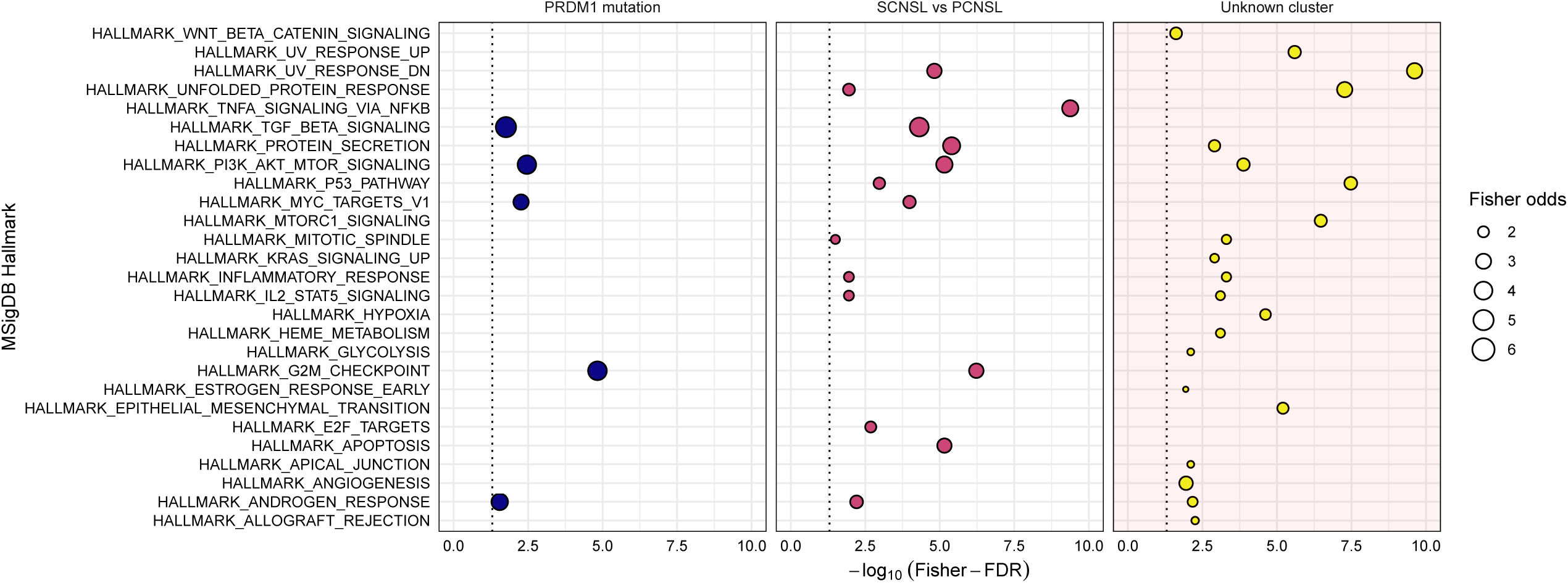
Gene set enrichment analysis. MSigDB pathways deregulated in specific comparisons, based on the differential expression patterns of miRNAs, from PRDM1 mutated and non-mutated or secondary and primary samples. Additionally, the figure shows the putatively deregulated pathways using the 19 miRNAs showing a significant association with the unknown “small cluster” based on the unsupervised clustering of expression data. The x axis shows the -log_10_ transformed, FDR corrected Fisher-test p-values, the y axis lists the deregulated MSigDB cancer hallmarks, and the dot size is proportional to the Fisher-test odds value. The dot colors correspond to the three different categories investigated (*PRDM1* mutated vs wild-type, SCNSL vs PCNSL and the unknown small cluster). The light red background of the “Unknown small cluster” facet indicates that pathways in this analysis are upregulated, while pathways in the other two comparisons are downregulated.

We carried out a similar analysis using the 19 miRNAs defined during the binary clustering, that are not expressed in our “small cluster”. However, in this case, as the miRNAs are not expressed in the small sample cluster, the associated enriched pathways are putatively upregulated in the same samples. Pathways include the unfolded protein response, connected to cellular stress, the P53 and MTORC1 signaling pathways, the epithelial to mesenchymal transition or the hypoxia gene sets and UV response genes.

Additional, more detailed results for the SCNSL vs PCNSL, *PRDM1* mutated vs non-mutated and “small cluster” analysis using the C2 – C7 gene sets are available in Supplementary file 6.

### miRNA expression profile association to survival characteristics

Finally, we asked whether the expression profile of specific miRNAs is associated to patient survival. We stratified patients according to the binary expression status of miRNAs and using the log-rank test, calculated the significance of overall survival (OS) differences between groups (Supplementary file 7) including both PCNSL and SCNSL samples. The only miRNA showing difference was hsa-miR-4488, with an FDR of 0.022 (Supplementary Figure 11 and 12) where expression of the miRNA was associated with worse survival. Median survival for patients with “Expressed” or “Not expressed” status of hsa-miR-4488 was one and 14 months, respectively. Repeating the analysis with only PCNSL samples (Supplementary file 7), we found no significant differences between groups after FDR correction. hsa-miR-18a-5p had the lowest FDR value (0.12). Of note, the survival analysis is hampered by the relatively low number of cases and the heterogeneous nature of the treatment regimens applied in this cohort.

### Validation of NanoString results

Based on the differential expression results, we selected a number of miRNAs for additional validation. We used quantitative RT-PCR and ddPCR for different miRNAs. (Supplementary file 8). Using the RT-PCR Δ-Ct values and the ddPCR counts, we correlated the validation results with the original and normalized NanoString read counts (Supplementary file 9). RT-PCR Δ-Ct values are expected to demonstrate a high negative correlation, while ddPCR counts are expected to show a high positive correlation with NanoString read counts or normalized expression values. We validated hsa-miR-411-5p and hsa-miR-32-5p using RT-PCR, where the correlation of Δ-Ct values with the normalized read counts was ≦ −0.6. Additionally, we validated hsa-let7g-5p, hsa-miR-191-5p and hsa-miR-379-5p using ddPCR where the correlation of ddPCR read counts with the NanoString read counts was ≧ 0.6, besides hsa-miR-411-5p where the correlation of normalized read count with ddPCR counts was 0.67 and the correlation of the original NanoString read count with the ddPCR count was 0.47.

## Discussion

CNS lymphomas represent a considerable clinical challenge, as their molecular pathogenesis is poorly explored. CNS lymphomas are difficult to investigate as they are rare, and usually small biopsies are obtained for diagnostic purposes, which may not be sufficient for further analyses [3, 4] in many cases. Moreover, the majority of these biopsies are archived as FFPE tissue blocks, which suffer from degradation of macromolecules [51-56]. However, miRNAs provide a robust signal and can be stably extracted from FFPE samples [20-22], making them ideal cancer biomarkers. They are also promising targets for molecular therapy in cancer, due to their role as oncomiRs or tumor-suppressors [57-59]. The NanoString nCounter technology allows direct quantitation of hundreds of miRNA transcripts from FFPE tissues with outstanding performance, therefore it may be a preferable choice over other transcriptomic methods [23-26].

Previous expression studies of PCNSL revealed various miRNAs with a potential diagnostic [28, 29, 31, 33, 34], prognostic [34-36] or predictive [33] value. These datasets were generated by different methods, and used diverse patient-derived sample types and control tissues [28-39], thus being difficult to synthesize and integrate. Moreover, most studies examined only a few miRNAs [28-30, 33, 34, 37] on a limited number of CNS lymphoma cases [28, 29, 32-39]. There is only a single study in the literature discussing the miRNA expression differences between PCNSL and SCNSL [31]. The authors demonstrated in CSF samples that miR-16, miR-30b, miR-30c, miR-191 and miR-204 were upregulated and miR-222 was downregulated in SCNSL, with miR-30c showing the largest expression difference [31].

In this study, we performed expression profiling of 798 human miRNAs in a large number of CNS lymphoma cases. We compared the miRNA expression patterns of FFPE brain biopsy specimens of primary and secondary CNS lymphomas to minimize the tissue bias that may well be presented with the examination of systemic (nodal) DLBCL. We used the NanoString platform, which is a reliable method for miRNA expression analysis of FFPE tissues [23-27].

We identified 28 up- and 3 downregulated miRNAs in SCNSL compared with PCNSL (Table 1). Out of these, 23 up- and all 3 downregulated miRNAs are completely novel and until now, were not described in the context of CNS lymphomas. Reassuringly, the remaining miRNAs described in the literature have similar expression patterns to our results. Amongst the already described miRNAs, we found miR-30c-5p, that is significantly increased in CSF samples of patients with SCNSL compared with PCNSL [31]. In general, miR-30c-5p has a tumor-suppressive role in cancer pathogenesis, and shows low expression in various malignancies (reviewed in [60]) which is in line with our findings (median expression: 11.18 and 0; rank: 137 and 147 in SCNSL and PCNSL samples, respectively) (Supplementary file 2). Its significant lower expression in PCNSL may contribute to their more aggressive behavior. Multiple studies have found higher expression of miR-21, a well-known oncomiR [61], both in PCNSL and DLBCL cases compared to controls [16, 28, 29, 33, 34, 62-66]. In our study, miR-21-5p generally showed high expression (median expression: 3278.55 and 2926.61; rank: 16 and 17 in SCNSL and PCNSL samples, respectively) with a significant increase in SCNSL cases. Higher expression of miR-21 has also been associated with worse overall survival in DLBCL patients [67]. We found 3 members of the miR-17-92 cluster (miR-19a-3p, miR-18a-5p and miR-106b-5p) to be upregulated in SCNSL compared with PCNSL, with a moderately high overall expression (median expression: 606.62 and 58.34, 1088.28 and 383.75, and 763.30 and 263.06; rank: 70 and 94, 52 and 72, and 62 and 75, respectively in SCNSL and PCNSL samples). This miRNA cluster has a strong oncogene activity in various malignancies including DLBCL [68-71]. Higher expression of miR-18a was found to be associated with a shorter overall survival in DLBCL [72]. Previous studies demonstrated other members of the miR-17-92 cluster (miR-17-5p and miR-20a) to be upregulated in PCNSL compared with nodal DLBCL [32, 37]. Moreover, high levels of miR-19b-1 and miR-92a-1 were detected in the CSF of PCNSL patients [28, 29].

Regarding the molecular subtypes, cases in the ABC group showed significantly higher expression of miR-155-5p, miR-222-3p and miR-522-3p and lower expression of miR-92a-3p compared with the GC group. In line with our results, higher expression of miR-155-5p [16, 19, 64, 65, 71, 73-76] and miR-222-3p [18, 71, 73] has already been associated with the ABC subtype in DLBCL. We found that the ABC molecular subtype also correlated with higher miR-522-3p, miR-454-3p and miR-455-5p expression compared with the UC subgroup.

This is the first study demonstrating differentially expressed miRNAs in association with the mutational status of the *PRDM1, MYC* and *CARD11* genes in CNS lymphomas. According to the literature, the only miRNA that has already been connected to any of these genes is miR-30a-5p, which directly targets *PRDM1* and modulates the WNT/beta-catenin pathway [77]. However, this association is not connected to the mutational status of *PRDM1* itself.

It is widely known, that the different miRNA profiling platforms do not perform consistently [24-27]. Nevertheless, we successfully validated the observed expression patterns of miR-148b-3p, miR-32-5p, miR-411-5p and miR-379-5p by ddPCR and/or RT-PCR methods.

Based on our data, pathway enrichment analysis revealed several downregulated pathways and gene sets in SCNSL compared with PCNSL. Additionally, *PRDM1* mutation was also associated with the downregulation of several pathways. Even though the evidence is circumstantial, as we did not directly measure the differential regulation of the genes comprising a pathway, but only their regulators, these pathways might be attractive targets for future drug development. The constitutive activation of NF-κB was already described in the literature [78-80] for DLBCL, apart from the activation of the PI3K-MTOR-AKT [79-81].

Based on our results, these pathways are generally more active in PCNSL compared to SCNSL and drugs [82] available to target these pathways might be more effective for a selection of PCNSL cases. The unfolded protein response pathway was similarly upregulated in PCNSL cases. This pathway is considered as a general pro-survival mechanism for cancer cells [83], and small molecule inhibitors targeting the pathway are becoming available, suggesting a possible therapeutic target in PCNSL, as these lymphomas might be more sensitive to treatment. Additional upregulated pathways in the PCNSL vs SCNSL comparison include protein secretion, the p53 pathway, MYC target genes, the G2M checkpoint, E2F transcription factor target genes, apoptosis genes, and genes related to androgen response. Considering that PCNSL is a rare and aggressive disease and the prognosis is poor [84], the pathways and molecular mechanisms analyzed in this study might be considered as novel drug targets. Considering the pathway level changes in *PRDM1* mutated samples, some drugs might be more effective in patients without *PRDM1* mutations. The TGF-Beta signaling, the PI3K-MTOR-AKT, MYC target genes, G2M checkpoint genes and androgen response genes are all downregulated in samples with *PRDM1* mutation, therefore the efficiency of their inhibitors might be decreased [80, 82, 85].

Intriguingly, principal component analysis (PCA) showed no clustering of cases according to either disease characteristics or subtypes even after accounting for the various batch effects and biases. Subsequent binary clustering of the cases according to miRNA expression (expressed or not expressed) revealed a small group of 8 samples clearly separating from the rest of the cohort. Additional sample clustering using a k-means based method with normalized expression values defined a sample cluster (SCluster1) consisting of 10 samples. Five samples from SCluster1 also overlapped with the small cluster defined in the binary expression analysis. Taken together, unsupervised clustering methods defined a small set of samples with markedly distinct miRNA expression patterns. Interestingly, the lack of expression of 19 miRNAs was found to be associated with the small cluster in the binary clustering analysis. Moreover, all of these 19 miRNAs were part of a miRNA k-means cluster (MCluster1) in SCluster1. Pathway enrichment analysis shows similar pattern in this small distinct set of samples to PCNSL, with even more pronounced changes compared to the SCNSL vs PCNSL analysis. The WNT/beta-catenin pathway activated here, was described as activated in PCNSL [86], and the small set of samples defined here might be a distinct PCNSL subgroup where the pathway can be efficiently targeted with Wnt inhibitors [87]. Based on these results, this is a well-defined sample group within the PCNSL cases contributing significantly to the distinct expression patterns between PCNSL and SCNSL.

Survival analysis of all cases using the binary expression data showed miR-4488 to be significantly associated with a worse overall survival, however, we did not find any association when analyzing PCNSL samples solely. It is important to highlight that these results are limited by the modest number of cases and the heterogeneous nature of the treatment regimens applied in this cohort.

## Conclusions

Our study identifies a novel CNS lymphoma subgroup defined by distinct miRNA expression patterns, describes putative subtype and cell-of-origin biomarkers, and proves the importance of specific miRNAs and pathways in CNS lymphoma pathogenesis. These results provide the basis for future research in the area of CNS lymphomas.

## Data Availability

The datasets supporting the conclusions of this article are included within the article, its additional files and at the NCBI GEO database with accession number GSE162956.

## Acknowledgements

We thank Rachel Bradshaw from NanoString Technologies for the microRNA profiling, Mingdong Liu from NanoString Technologies for help in NanoString data QC and analysis, and Zoltán Szállási for critical reading of the manuscript.

## Declarations

### Ethics approval and consent to participate

Permissions to use the archived tissue have been obtained from the Local Ethical Committee (TUKEB-1552012) and from BRAIN UK (Ref.: 16/018), and the study was conducted in accordance with the Declaration of Helsinki.

### Consent for publication

Not applicable.

