## Supplementary file 10 for "Distinct microRNA expression signatures of primary and secondary central nervous system lymphomas"

**Supplementary files**

The following supplementary files are available for the manuscript.

**Supplementary file 1.** Sample information. The table contains available clinical and sample information including age, sex, survival status, clinical treatment, mutation status of selected genes, tumor content, primary/secondary and molecular subtype classification, sample and RNA isolation year, institute, RNA isolation group, and scan date.

**Supplementary file 2.** Summary statistics of normalized NanoString read counts. The first tab includes the minimum (*expression_min*), maximum (*expression_max*) and average expression (*expression_mean*) across all samples, together with the number of samples where the miRNA was expressed (*is_expressed*). A miRNA was considered expressed if it had a higher expression than the lowest positive control. The second tab contains minimum (*expression_min*), maximum (*expression_max*), average (*expression_mean*), and median (*expression_median*) expression across PCNSL or SCNSL samples (*pri_sec*). Additionally, it contains the rank of median expression among all miRNAs (*expression_rank*) and the expression groups (*group*), based on the ranks.

**Supplementary file 3.** Differential expression results. Limma-voom based differential expression results. The table contains the log2 fold change (*log2fc*), and false discovery rate (*FDR*) for each miRNA in each comparison (*Condition*). Additionally, it contains the number of samples with a specific mutation (*Mutation_count*), for the mutated – non-mutated sample comparisons and a Yes/No categorization showing if a specific miRNA was considered changing significantly in a specific condition (*significant*).

**Supplementary file 4.** Sample clusters. Summary of different clustering analysis methods, showing sample ids (*sample_id*) that were included in the unknown "small cluster" based on a specific method (*Binary cluster PCNSL+SCNSL*, *Binary cluster PCNSL only*, *Kmeans cluster PCNSL+SCNSL*, *Kmeans cluster PCNSL only*). The table shows the number of times a sample id was present in the small cluster during a specific analysis (*Present in cluster*) and the size of the cluster in a specific analysis (*Cluster size*).

**Supplementary file 5.** Small cluster associated miRNAs. List of miRNAs that show a significant association with the "small cluster". The table shows the miRNA id (*mirna_id*), the number of samples in the two clusters with expressed/not expressed status (*Big_exp*, *Small_exp*, *Big_noexp*, *Small_noexp*), the Fisher-test p-value (*fisher_p*), odds ratio (*fisher_odds*) and FDR (*fisher_fdr*).

**Supplementary file 6.** Pathway enrichment analysis. The table shows all MSigDB pathways and gene collections, that were significantly enriched in a specific condition. Besides the condition (*condition*) and the MSigDB id (*pathway*), it includes the MSigDB class (*msigdb_class*), the number of genes considered differentially expressed/not differentially expressed, in the pathway/not in the pathway (*pathway_de*, *pathway_not_de*, *not_pathway_de*, *not_pathway_not_de*), the Fisher-test p-value (*fisher_p*), odds ratio (*fisher_odds*) and FDR (*fisher_fdr*).

**Supplementary file 7.** Survival analysis. The table shows the results of the survival analysis, where samples were stratified based on the expressed/not expressed status of a miRNA. Columns include the miRNA id (*mirna_id*), log-rank survival test p-value (*pval*), FDR (*FDR*), the number of samples with or without expression (*n0* and *n1*) and the survival analysis category (*category*).

**Supplementary file 8.** Validation results. Results of the RT-PCR and ddPCR validations, including miRNA id (*mirna_id*), sample id (*sample_id*), box number (*bx_no*), validation type (*measurement*), Ct, Δ-Ct, or ddPCR count values (*value*).

**Supplementary file 9.** Correlation of PCR based validation and NanoString data. Spearman correlation of RT-PCR Δ-Ct or ddPCR (*condition*) count values with the NanoString counts (*spearman_corr_count*) or normalized (*spearman_corr_expr*) NanoString values for the validated miRNAs (*mirna_id*), besides the number of samples (*sample_number*) used for validation.

**Supplementary figure**

The following supplementary figures are available for the manuscript.

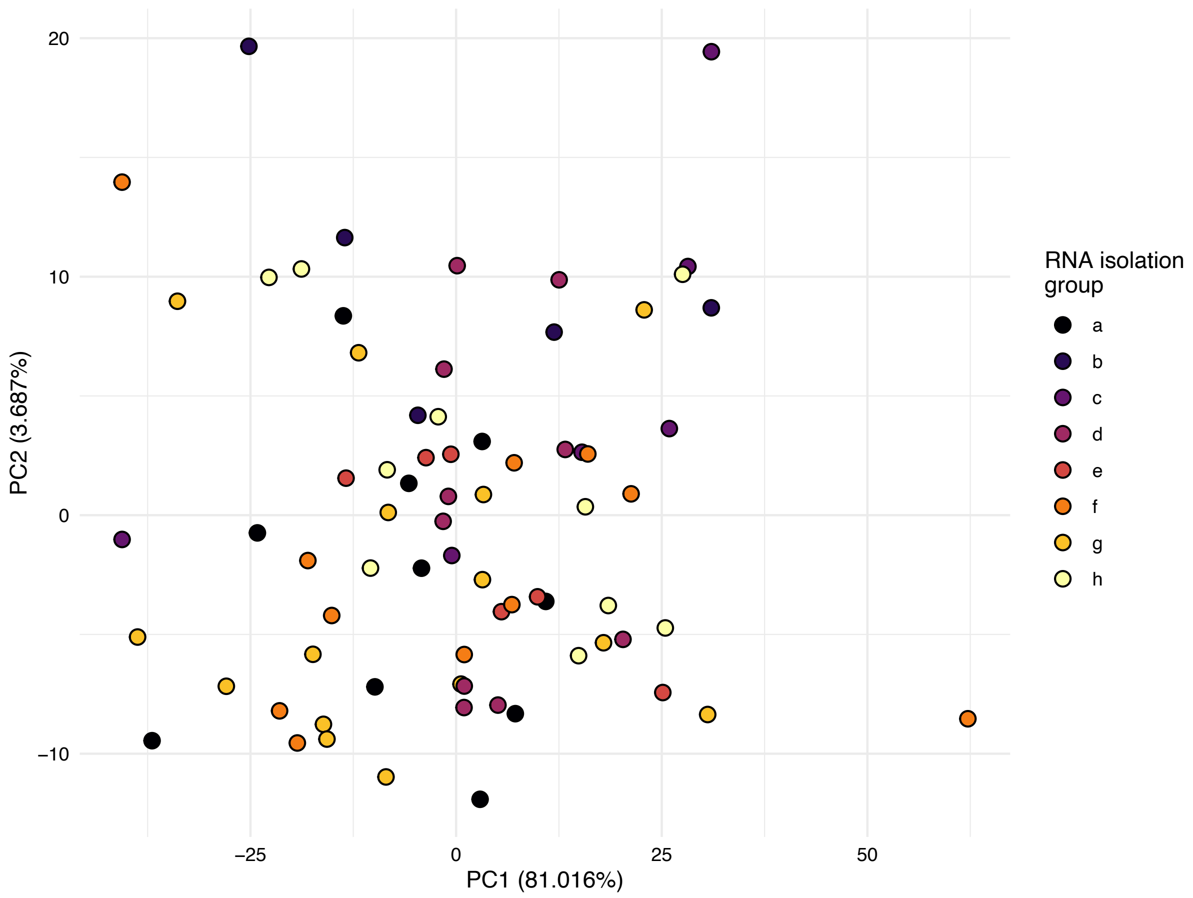


**Supplementary figure 1.**

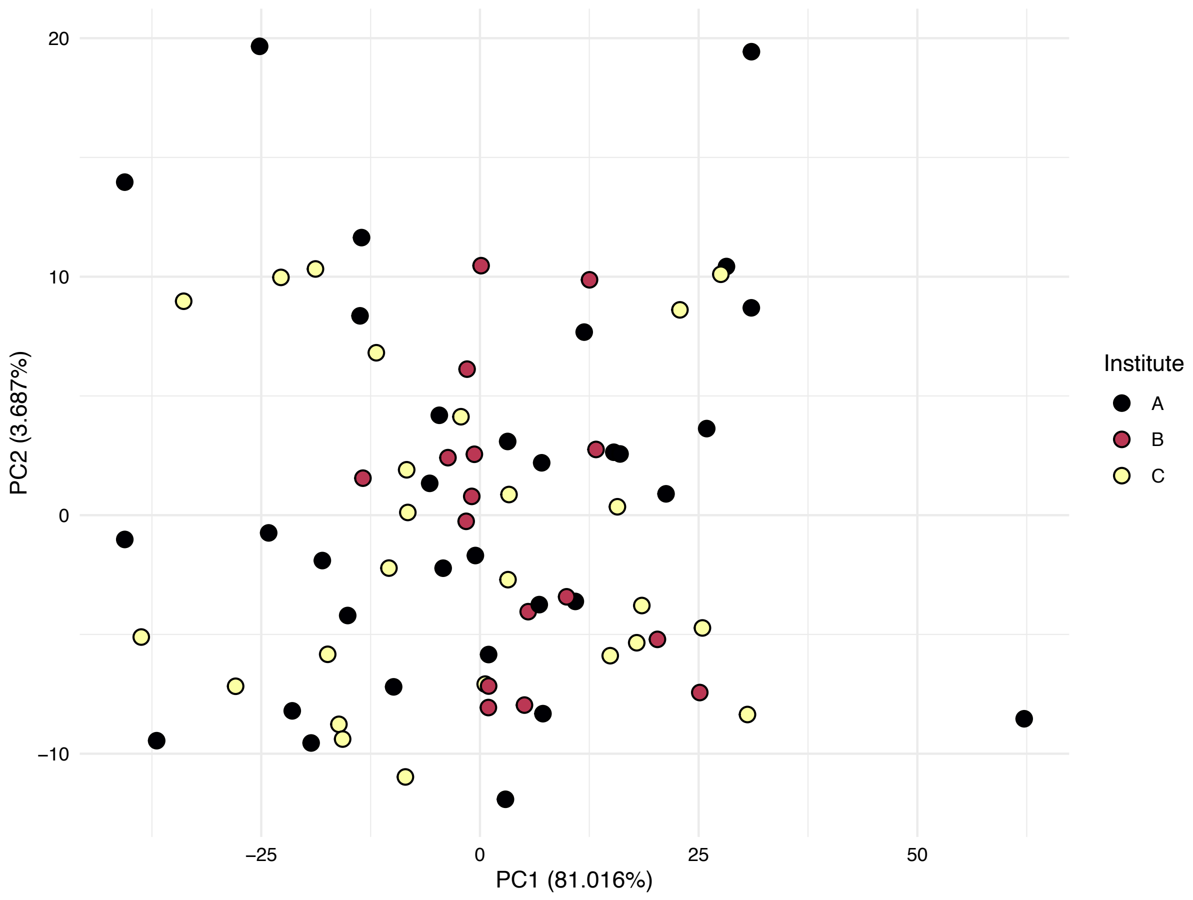


**Supplementary figure 2.**

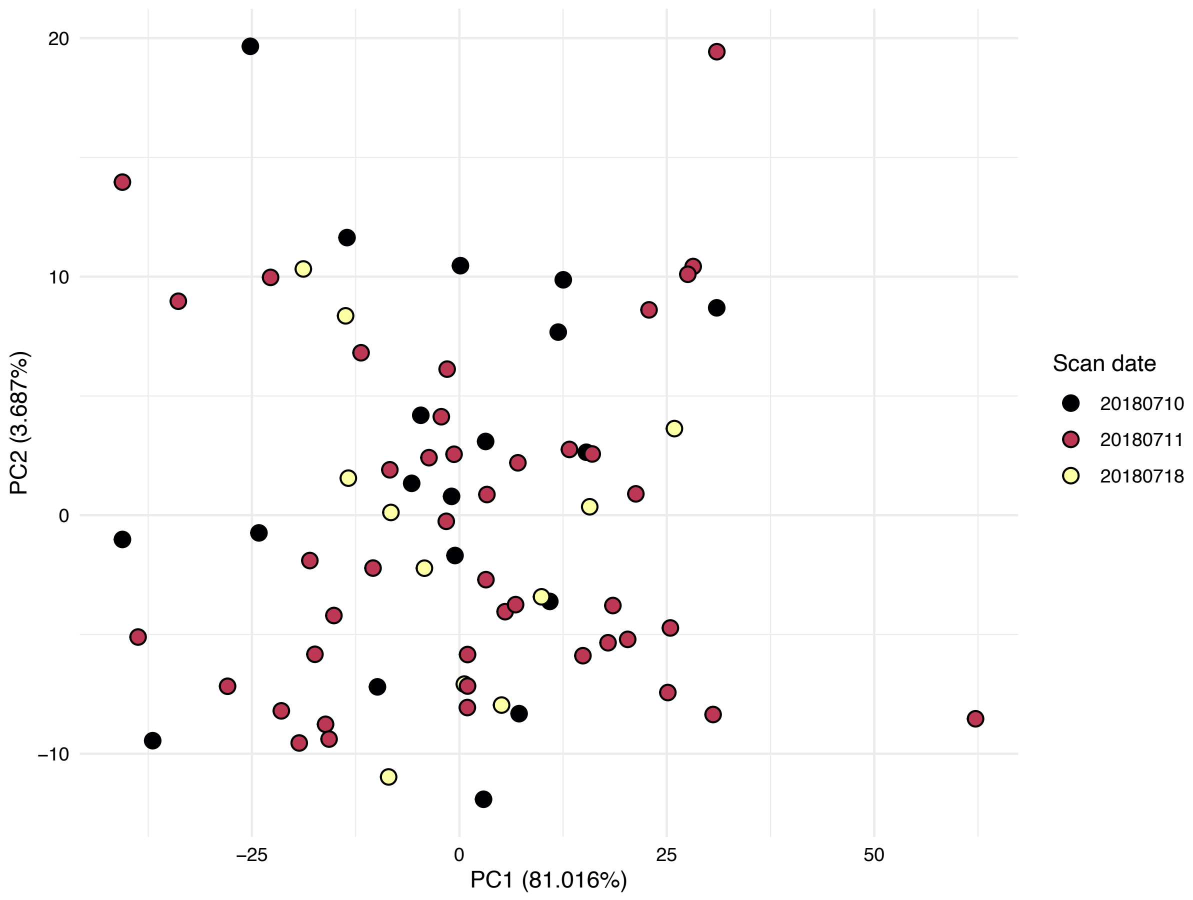


**Supplementary figure 3.**

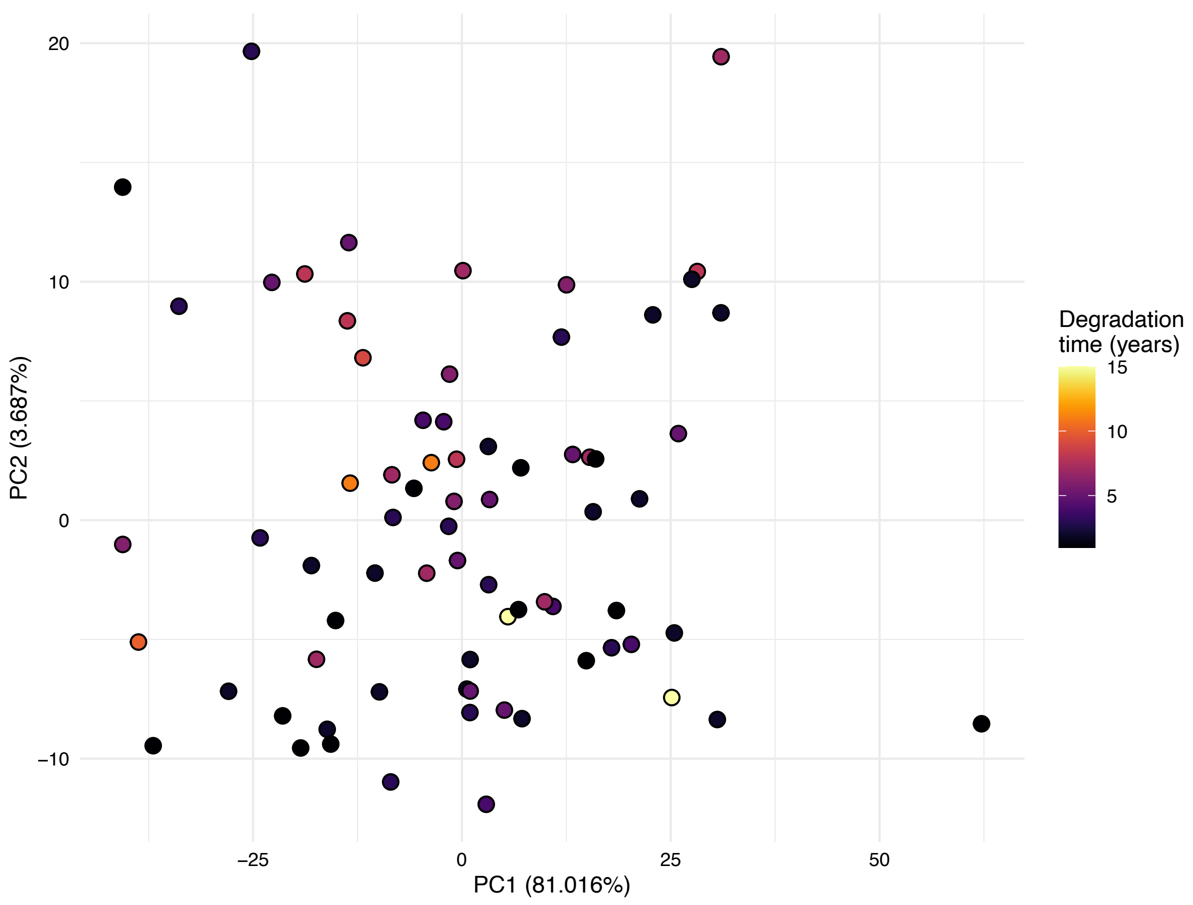


**Supplementary figure 4.**

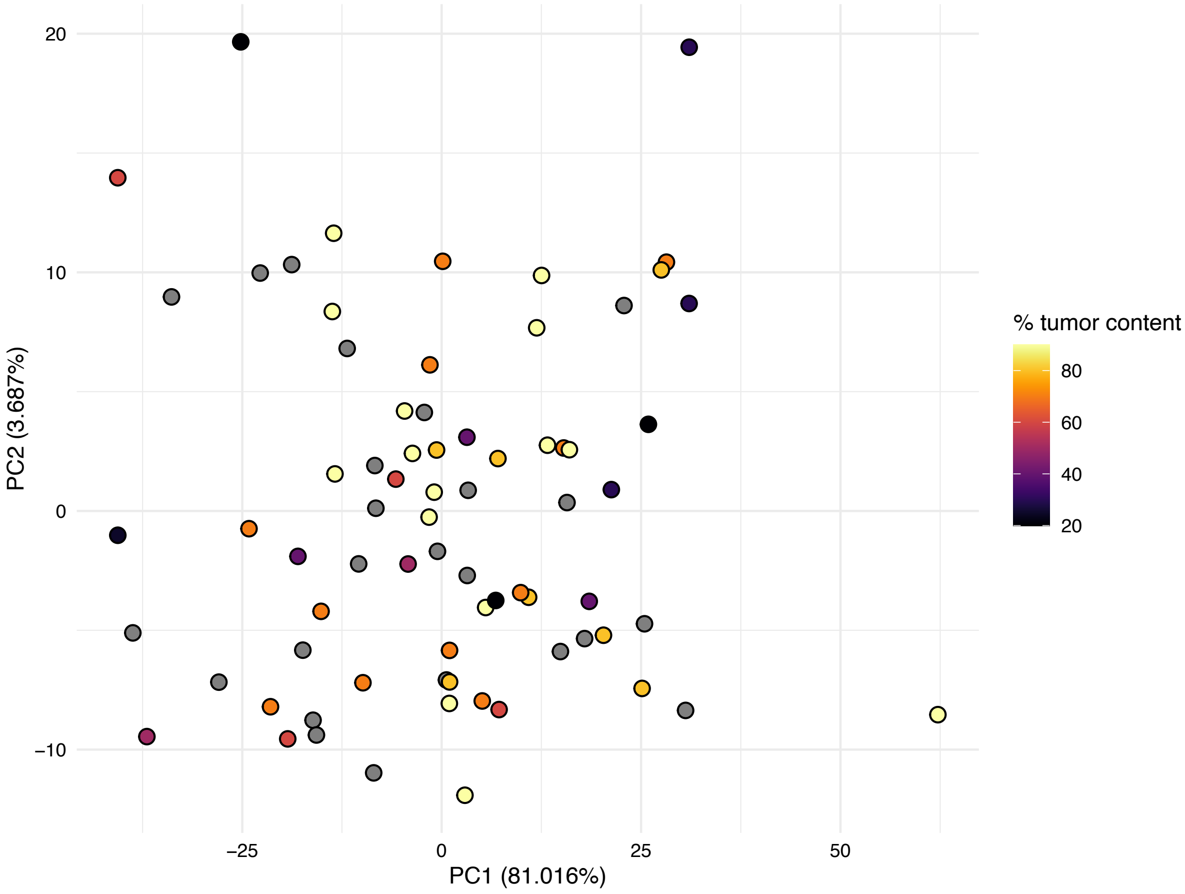


**Supplementary figure 5.**

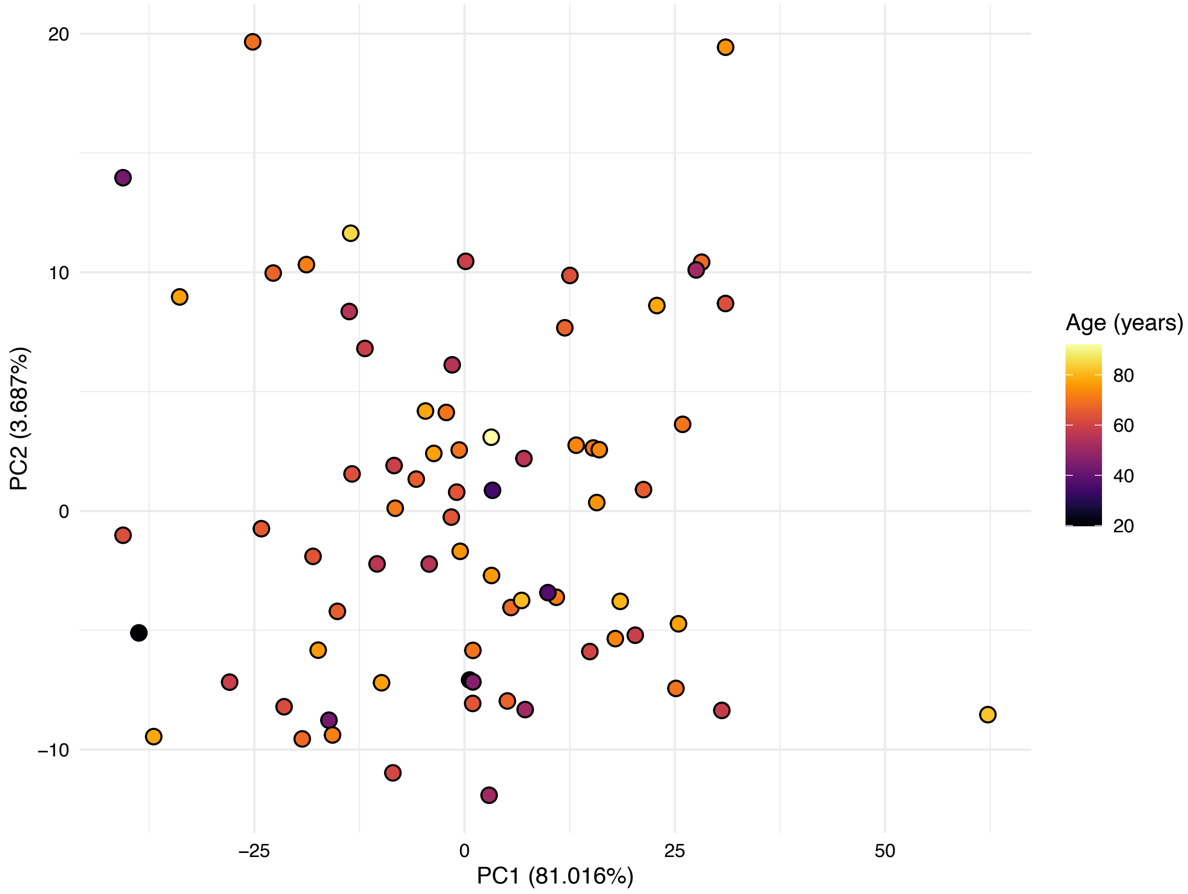


**Supplementary figure 6.**

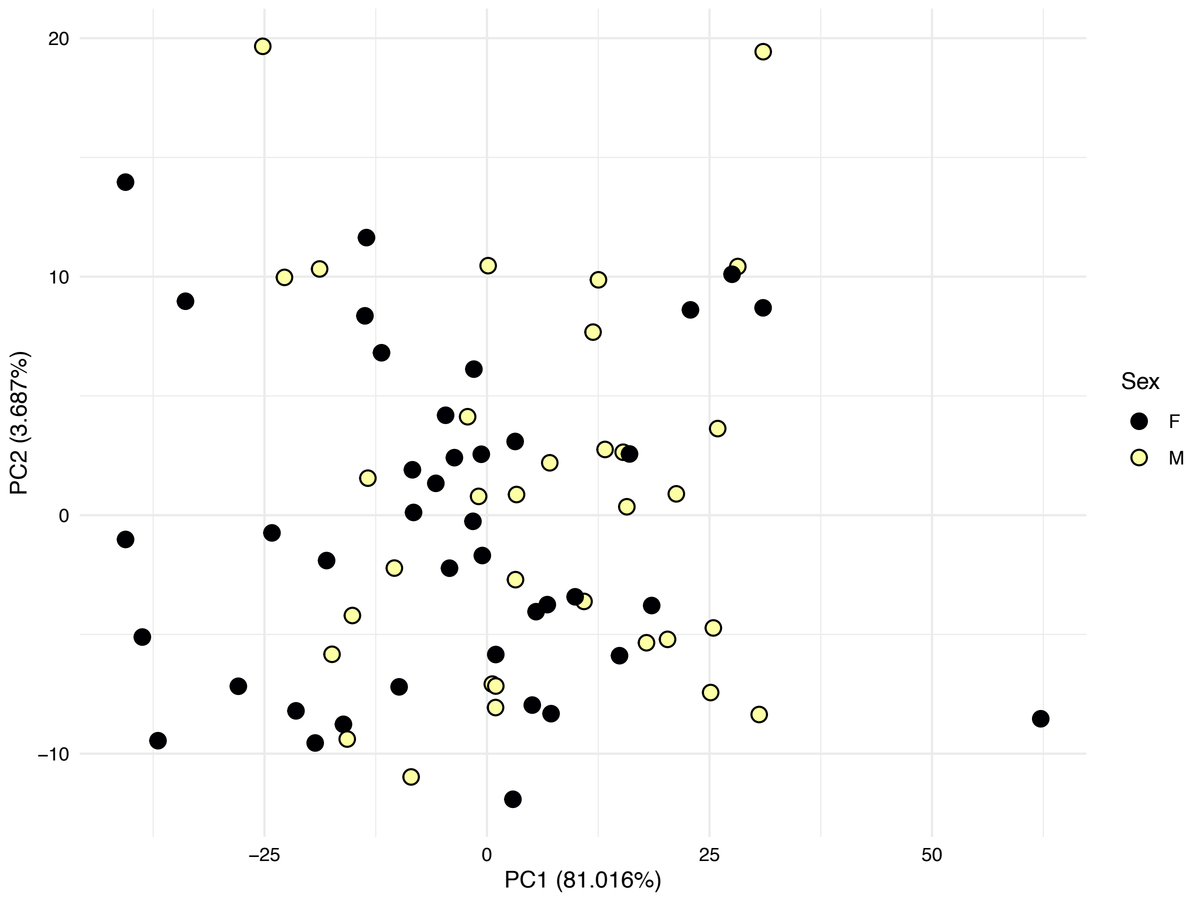


**Supplementary figure 7.**

**Supplementary figure 1-7.** Principal component analysis of miRNA expression patterns of all samples, after removing potential batch effects from the normalized and voom transformed data. The X and Y axes show the first two principal components, with the % variance explained in parenthesis. The color of the points corresponds to various possible technical and biological confounding factors, including RNA isolation group, institute, scan date, degradation time in years, % tumor content, age or sex. Grey points correspond to samples with missing information.

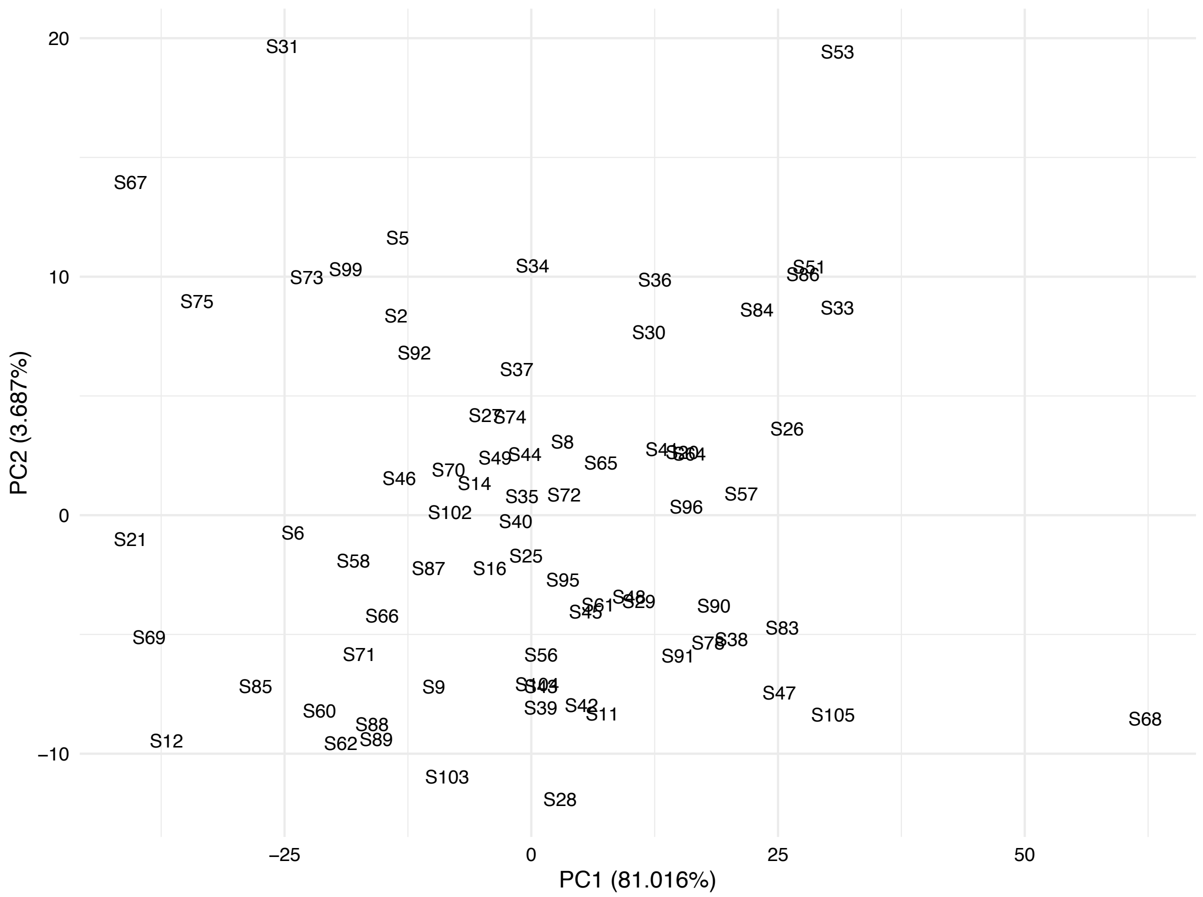


**Supplementary figure 8.** Principal component analysis of miRNA expression patterns of all samples, after removing potential batch effects from the normalized and voom transformed data. The plot shows the sample IDs. The X and Y axes show the first two principal components, with the % variance explained in parenthesis.

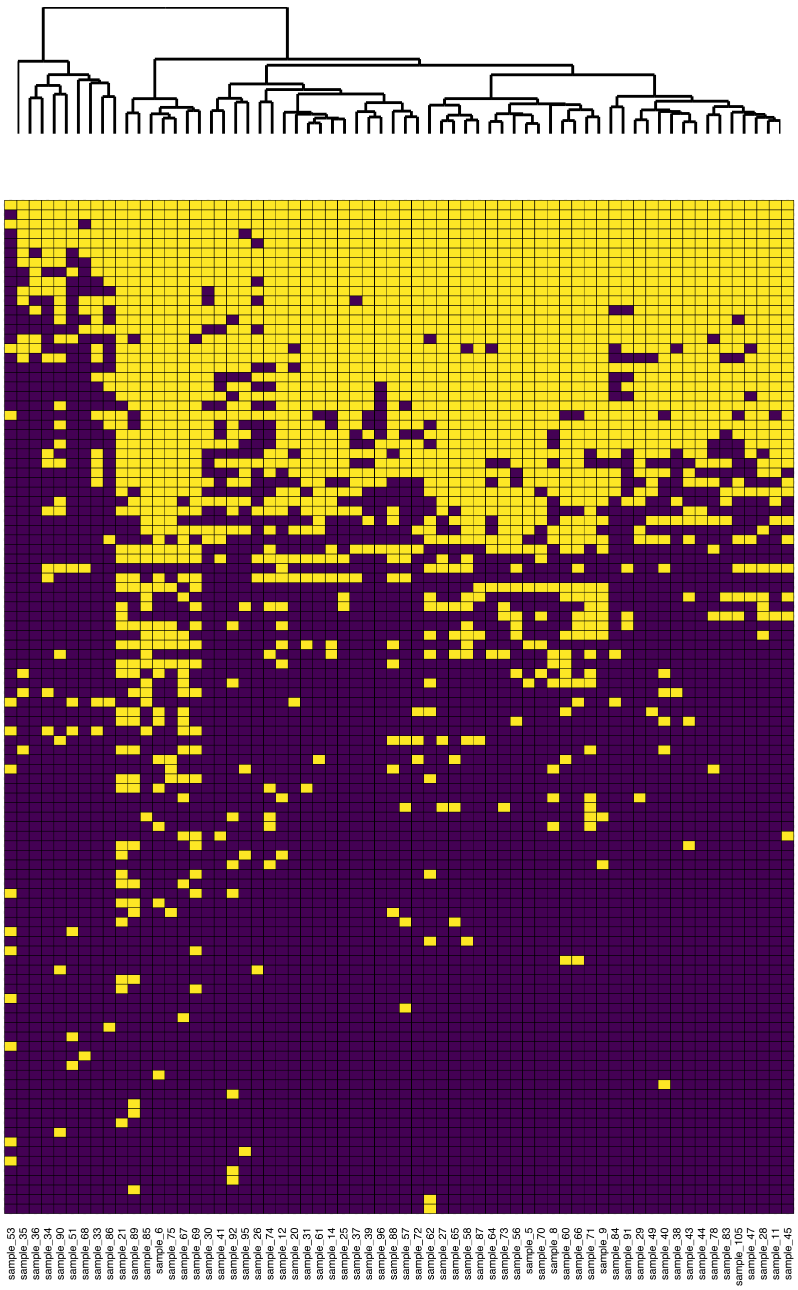


**Supplementary figure 9.** The unsupervised clustering of samples, similar to **Figure 3.**, but excluding SCNSL samples.

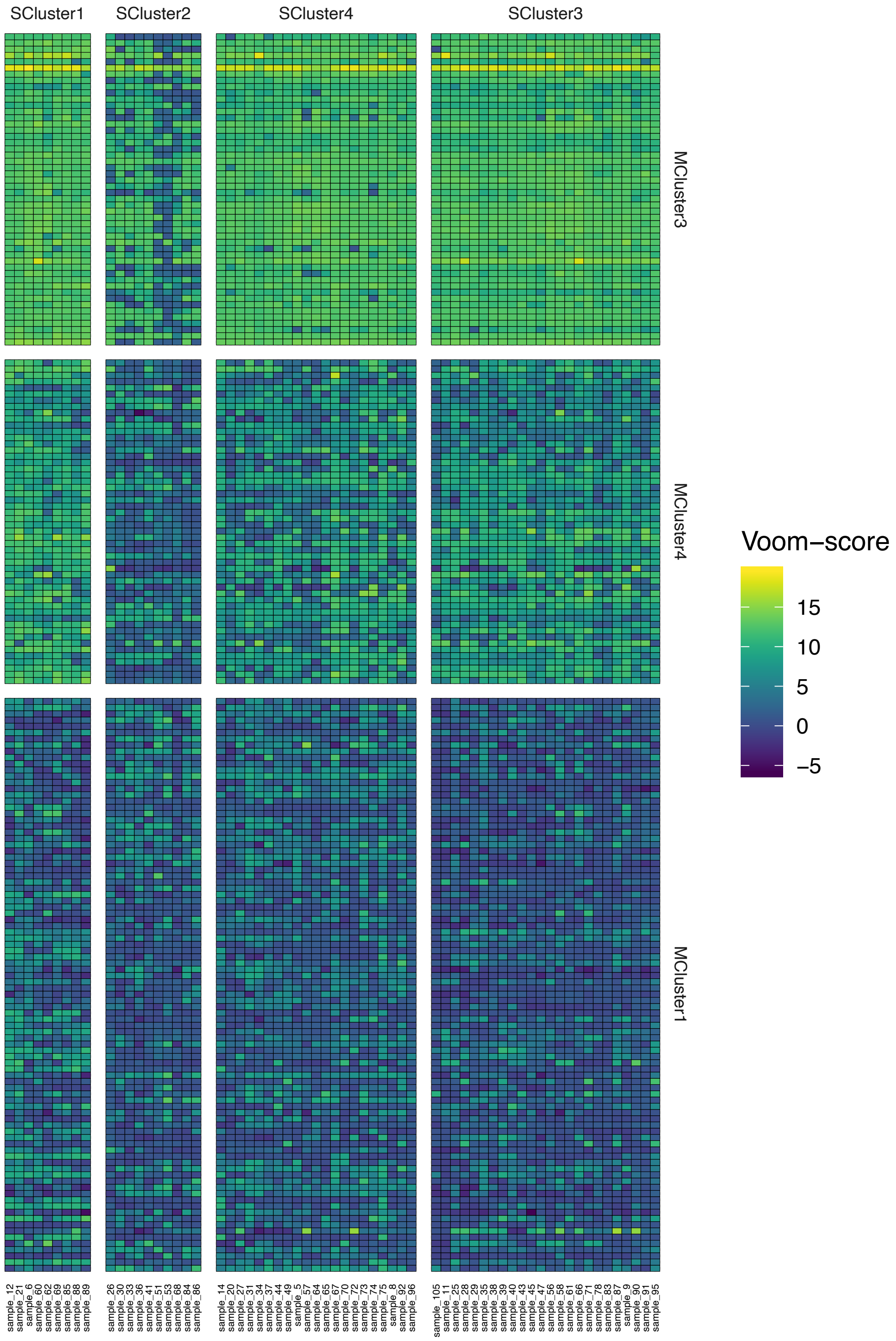


**Supplementary figure 10.** The unsupervised clustering of samples using normalized miRNA expression patterns, similar to Figure 4., but excluding SCNSL samples.

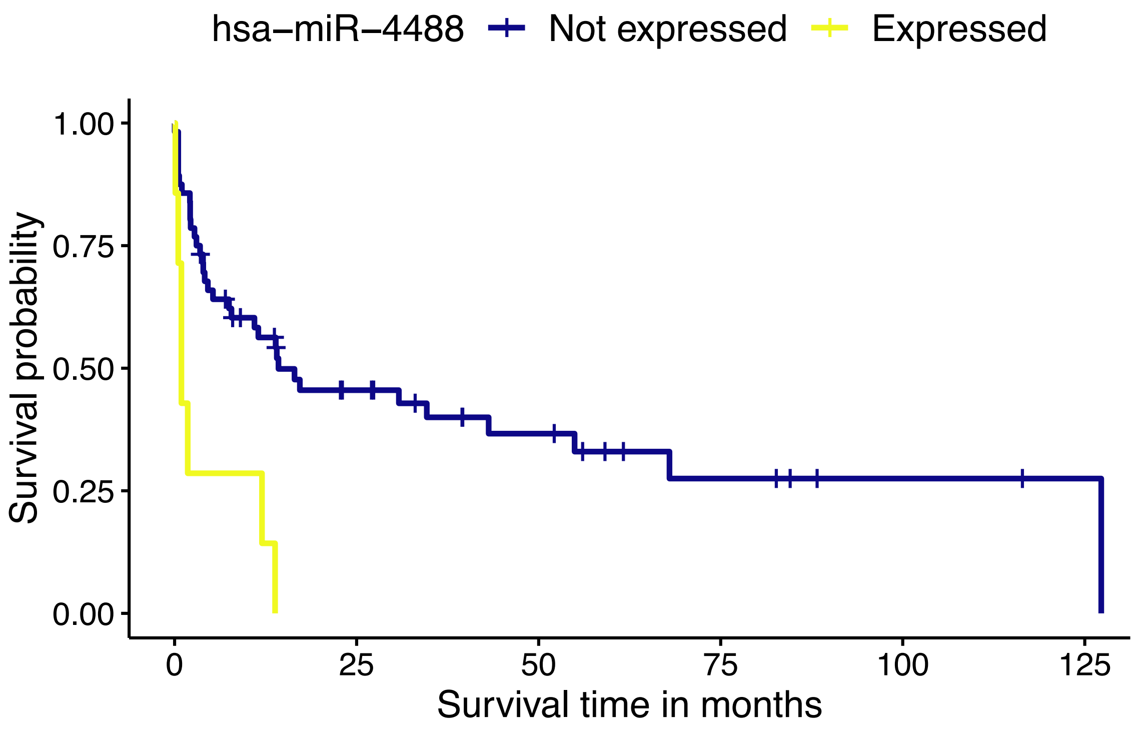


**Supplementary figure 11.** Survival curve of patients stratified based on the binary expression (ON/OFF) of hsa-miR-4488.

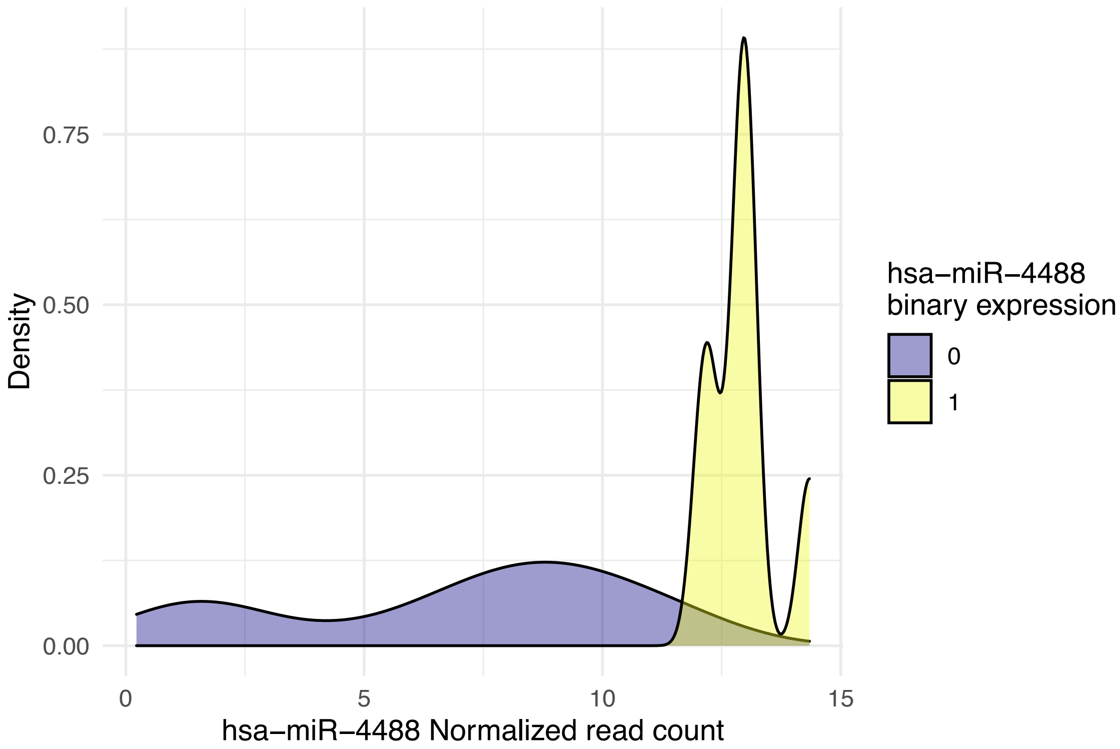


**Supplementary figure 12.** Normalized NanoString read count density of hsa-miR-4488 across samples, separated by the binary expression (ON/OFF) classification. The hsa-miR-4488 miRNA in a specific sample might be considered not expressed, even with a larger than zero number of reads, if it is smaller than the read count of the smallest positive control.
